# Whole genome analysis reveals the genomic complexity in metastatic cutaneous squamous cell carcinoma

**DOI:** 10.1101/2022.01.10.22269035

**Authors:** Amarinder S. Thind, Bruce Ashford, Dario Strbenac, Ruta Gupta, Jonathan R Clark, N. Gopalakrishna Iyer, Jenny Mitchell, Jenny Lee, Simon A Mueller, Elahe Minaei, Jay R. Perry, Marie Ranson

**Affiliations:** School of Medicine, University of Wollongong, Wollongong, NSW, Australia; Illawarra Health and Medical Research Institute, Wollongong, NSW, Australia; Illawarra Shoalhaven Local Health District Wollongong, NSW, Australia; Sydney Medical School, Faculty of Medicine and Health, The University of Sydney, NSW, Australia; Anatomical Pathology, Royal Prince Alfred Hospital, Sydney, NSW, Australia; Royal Prince Alfred Institute of Academic Surgery, Sydney Local Health District, Sydney, NSW, Australia; Department of Head and Neck Surgery, Chris O’Brien Lifehouse, Sydney, NSW, Australia; National Cancer Center, Singapore; Department of Medical Oncology, Chris O’Brien Lifehouse, Sydney, NSW, Australia; Department of Clinical Medicine, Macquarie University, Sydney, NSW, Australia; Department of Otorhinolaryngology, Head and Neck Surgery, Zurich University Hospital and University of Zurich, Zurich, Switzerland; School of Chemistry and Molecular Bioscience, University of Wollongong, Wollongong, NSW, Australia

**Keywords:** whole genome sequencing, cutaneous squamous cell carcinoma, metastases, non-coding mutations, UTR, cSCC

## Abstract

Metastatic cutaneous squamous cell carcinoma (cSCC) is a highly morbid disease requiring radical surgery and adjuvant therapy that is associated with reduced overall survival. Yet compared to other advanced malignancies, relatively little is known of the genomic landscape of metastatic cSCC. We have previously reported the mutational signatures and mutational patterns of CCCTC-binding factor (CTCF) regions in metastatic cSCC. However, many other genomic components (indel signatures, non-coding drivers, and structural variants) of metastatic cSCC have not been reported. To this end, we performed whole genome sequencing on lymph node metastases and blood DNA from 25 cSCC patients with regional metastases of the head and neck. We designed a multifaceted computational analysis at the whole genome level to provide a more comprehensive perspective of the genomic landscape of metastatic cSCC.

In the noncoding genome, 3’UTR regions of *EVC* (48% of specimens), *PPP1R1A* (48% of specimens) and *ABCA4* (20% of specimens) along with the tumor-suppressing lncRNA LINC01003 (64% of specimens) were significantly functionally altered (Q-value < 0.05) and represent potential noncoding biomarkers of cSCC. Recurrent copy number loss in the tumor suppressor gene *PTPRD* was observed. Gene amplification was much less frequent and few genes were recurrently amplified. Single nucleotide variants driver analyses from 3 tools confirmed *TP53* and *CDKN2A* as recurrently mutated genes but also identified *C9* as potential novel driver in this disease. Further, indel signature analysis highlighted the dominance of ID signature 13 (ID13) followed by ID8 and ID9. ID 9 has previously been shown to have no association with skin melanoma, unlike ID 13 and 8, suggesting a novel pattern of indel variation in metastatic cSCC. The enrichment analysis of various genetically altered candidates shows enrichment of ‘TGF-beta regulation of extracellular matrix’ and ‘Cell cycle G1 to S check points’. These enriched terms are associated with genetic instability, cell proliferation, and migration providing mechanisms of genomic drivers of metastatic cSCC.

## INTRODUCTION

Cutaneous squamous cell carcinoma (cSCC) is the second most common malignancy, after basal cell carcinoma (BCC), affecting up to 1 000 000 people in the United States (US) annually [1]. In time, and as a result of the ageing population and changing ratios of BCC/cSCC, the mortality rate of cSCC is likely to exceed that of melanoma [2]. Although primary cSCC is common, metastasis only occurs in 2-5% of cSCC [3-5]. cSCC arising in the head and neck generally show a predictable pattern of spread, predominantly metastasizing to the intraparotid, level II (upper jugular) and perifacial lymph nodes [4]. cSCC that have metastasized to regional lymph nodes are associated with a worse prognosis [6], with modest progress made in the management of regionally advanced disease over the last 15 years. Most patients with regional metastases from cSCC of the head and neck are managed with a multimodality approach, which usually involves surgery (parotidectomy and neck dissection) and adjuvant external beam radiotherapy depending on the site and stage at the time of diagnosis [7-9]. More recently immunotherapy has attracted great interest as a potential alternative for unresectable or distant metastatic disease [10, 11].

Despite the very high incidence, relatively little is known regarding the genomic landscape of metastatic cSCC. We have previously described the genomic mutational burden, mutational signatures, and mutations in CCCTC-binding factor regions using whole genome sequencing (WGS) data from 15 cSCC metastases [12]. However, the majority of studies to date have reported on somatic variation in primary cSCC [13-16] and/or cSCC metastases [16-20], using whole exome sequencing (WES) and/or targeted next generation sequencing, which by definition focusses on the coding genome. Thus, the extent of analysis of non-coding (including regulatory) regions of the genome is limited and varies across studies. Pickering et al [20], the only study employing WES, and incorporating 32 primary and only 7 metastatic samples, did not include regulatory or non-exome regions analysis. Both Li et. al [18] (29 lymph node metastatic formalin fixed paraffin embedded (FFPE) samples) and Zehir et. al [17] (MSK-IMPACT) (28 primary and 27 metastatic FFPE samples) used targeted NGS, with limited non-coding analysis. Zehir et al [17] specifically included the *TERT* promoter in their targeted panels but otherwise included no regulatory elements. Yilmaz et al. [16] performed WES and/or targeted NGS on 18 metastatic and 10 primary FFPE cSCC samples and reported coding gene drivers based purely on mutational frequencies, without adjusting for gene length or covariates. Additional functional driver predictions analysis would be required to confidently call genes as drivers [21]. Furthermore, FFPE processing has well-known impacts on the quality of DNA for sequencing analyses [22] and it is important to note that for most of the metastatic studies samples are collected using FFPE. Li et al [18] similarly did not include regulatory or non-coding variant analysis. Furthermore, none of these studies addressed variation in either 5’ or 3’ untranslated regions (UTR) or other non-coding elements such as promoters (other than *TERT* promoter) or long non-coding RNAs. Sequence variants occurring within these functional non-coding elements are important as they have the potential to alter gene expression. For example, lncRNA are thought to influence expression of proteins by pre- and post-translational influences on DNA/RNA and proteins, chromatin function, miRNA activity and signaling pathways by an array of mechanisms [23, 24]. 3’UTRs regulate crucial aspects of post-transcriptional gene regulation [25]. Mutations in these regions can deregulate gene expression by disrupting miRNA-mRNA interactions, which in both tumor suppressor genes and oncogenes can drive cancer progression [26, 27]. This variation in so called *cis-elements* can also impact gene expression by altering translation initiation in cancer [28].

Given the shortcomings associated with WES and NGS analyses of complex genomes, in the current report we have performed WGS on 25 metastatic cSCC samples and applied a detailed, multifaceted computational analysis at the whole genome level to provide a comprehensive understanding of the genomic landscape of metastatic cSCC. This included processing of WGS data for somatic variations in both coding and non-coding regions, and indel signatures, apart from structural variants and copy number alterations analyses. For non-coding genomic regions, we have focussed on UTRs, lncRNA and promoters regions as these represent non-coding regions that are most accessible to interrogation in high mutational burden tumours using currently available tools.

## MATERIALS AND METHODS

### Study population, sample collection and processing

This study was undertaken with Institutional Human Research Ethics approval (UOW/ISLHD HREC14/397). Thirty-two patients with resectable metastatic cSCC (28 from males and 4 from females) were identified by the treating surgeons preoperatively. In addition to whole blood (for germline DNA), sections of fresh tumor from nodal metastases were collected during surgery and immediately snap frozen. These sections were used for both DNA/RNA extraction (Qiagen AllPrep, Qiagen, Hilden, Germany) and for cellularity estimates. Only samples with > 30% tumor (range 35-95%) proceeded to DNA quality control (QC). QC comprised spectrophotometry (Nanodrop 2000 ThermoFisher Scientific Inc), gel-electrophoresis and SNP array. Of the 32 samples sequenced, 25 passed QC (96% from males) (Table 1). The remaining 7 samples had insufficient clonal tumor content (median variant reads ≤ 5 or median VAF < 0.1) or had an extreme GC bias as determined by PURPLE [29]. Briefly, if more than 220 copy number segments were unsupported by a corresponding structural variants at either end, the sample was flagged as Fail-Segment. The mean sequencing coverage of the 25 samples was 94.56× (range: 64-143) for tumor and 41.08× (range: 30-56) for blood.

**Table 1.** Clinicopathologic data of the cohort of 25 patients with cSCC lymph node metastases.

| **Sample** | **Primary location** | **Metastasis location** | **Immuno-suppressive treatment** |
| --- | --- | --- | --- |
| CSCC_0001 | left lip | left neck | no |
| CSCC_0002 | right ear | right parotid | no |
| CSCC_0003 | unknown | right parotid | no |
| CSCC_0004 | bilateral lip | bilateral neck | no |
| CSCC_0005 | left forehead | left parotid | no |
| CSCC_0006 | left cheek | left neck | azathioprine |
| CSCC_0007 | unknown | left neck | no |
| CSCC_0009 | bilateral forehead | right neck | cyclosporine A, tacrolimus |
| CSCC_0010 | left scalp | left neck | no |
| CSCC_0011 | unknown | right parotid | no |
| CSCC_0012 | right nose | right neck | no |
| CSCC_0013 | right pinna | right parotid | no |
| CSCC_0014 | left cheek | left perifacial | no |
| CSCC_0022 | scalp | left neck | no |
| CSCC_0024 | lip | right neck | no |
| CSCC_0025 | parotid | Parotid | no |
| CSCC_0066 | Unknown | Parotid | no |
| CSCC_0124 | Parotid | Parotid | no |
| CSCC_0125 | parotid | parotid | no |
| CSCC_0126 | left temple | left neck | no |
| CSCC_0130 | unknown | left parotid | no |
| CSCC_0132 | right ear | parotid/neck | no |
| CSCC_0133 | unknown | parotid | no |
| CSCC_0134 | unknown | right neck | no |
| CSCC_0135 | unknown | right neck | no |

### Variant calling and functional significance of SNVs and indels

FASTQ reads were aligned to reference genome GRChr38 using BWA-kit version 0.7.17 (BWA-MEM read aligner) (for details refer to https://github.com/Sydney-Informatics-Hub/Fastq-to-BAM). The Genome Analysis Tool Kit (GATK) 4.1.2.0 and its BaseRecalibrator tool was used to refine the read alignment. Single nucleotide polymorphisms (SNPs) and insertion-deletion (indel) variants were called by implementing GATK’s Best Practices Workflow. These pipelines use HaplotypeCaller for germline short variant discovery and Mutech2 caller for somatic short variant discovery for SNVs and Indels (for details refer to https://github.com/Sydney-Informatics-Hub/Somatic-ShortV). Furthermore, variants effect prediction and annotations were completed using OpenCravat platform [30]. Mutation Annotation Format (MAF) files were generated based with Variant Effect Predictor annotations. Three different methods for driver discovery were then used; OncodriveFML[31], MutSigCV [21] and dNdScv [32].

OncodriveFML predicts the functional significance of both coding and non-coding variants as it is one of the few tools designed for non-coding genomic analysis [31]. It first determines the functional impact of the observed somatic mutations using Combined Annotation Dependent Depletion (CADD) for specified genomic elements (UTR, promotor, coding regions) across the cohort. Later, for the statistical significance, it compares the average functional impact score of the observed mutations in the element with the average functional impact scores of a similar number of the random mutational set. The CADD score provides a priority for identifying mutations with functional, deleterious, and pathogenic impacts. These scores are calculated by combining the information from multiple annotations into a single metric.

MutSigCV identifies genes that are mutated more often than expected by chance and reduces the number of false positives in the generated list of significant genes, which is especially useful for tumors, such as metastatic cSCC, with high mutation rates [21]. This is achieved by incorporating various types of information such as patient-specific mutation frequencies and mutation spectra, gene-specific mutation rates, expression levels and replication times.

dNdScv is designed to test for positive and negative selection in cancer genomes [32]. As UV-induced cancer genomes such as cSCC can affect the accuracy of the dNdScv model we carefully monitored the annotation of CC>TT changes (sometimes reported as C>T changes). Results report significance for missense and truncating mutations, as well as indels as global p-values. Genes that were falsely flagged as significant with negative selection were not considered for this analysis.

For downstream analysis, genes predicted to be driver genes by at least two of these tools were considered. Firstly, genes with significance p-values <0.005 were filtered from each of the 3 tools, and shared genes determined using a Venn diagram. We then compared the functional impact of SNVs in these selected driver genes to previously reported primary and metastatic cSCC data [17, 18, 20, 33] available on cBioportal [34]. This included 92 samples of nodal metastatic cSCC (WES= 10, targeted NGS = 82) and 88 samples of primary cSCC (WES=32, targeted NGS=56).

### Copy Number Variation

Copy number alterations in the 25 metastatic genomes was derived using PURity & PLoidy Estimator (PURPLE) [29], which estimates copy number and purity of tumor sample by using read depth ratio from COBALT and tumor B-allele frequency (BAF) from AMBER. The pipeline is available at github of HMF Tools (https://github.com/hartwigmedical/hmftools). Driver genes with significant amplifications and deletions were then identified using PURPLE driver copy number outputs. For driver genes, PURPLE searches for genes with high level amplification (minimum Exonic Copy number > 3 * sample ploidy) and deletion (minimum exonic copy number< 0.5) and then uses iteration to establish the most significant focal peaks.

GRIDSS2 and its companion interpreter tool LINX were employed for somatic structural variant analysis and Gene fusion [35]. COSMIC3 based SNVs and Indels signatures from the whole genome were built using MutationalPatterns [36] software, respectively.

The driver gene candidates obtained from various genetic alteration analyses such as copy number variation drivers, somatic variant drivers, and other non-coding drivers were combined for enrichment analysis. In the case of copy number gain/loss, we selected only those genes affected in >20% of the samples in our cohort. Using the Enrichr web application [37] we determined the involvement of the candidate driver genes in various cellular components of the cells, biological pathways and predicted miRNA and drug targets.

## RESULTS

### Patient characteristics and clinicopathologic data

Twenty-five metastatic cSCC samples from lymph nodes in the head and neck region were collected between 2015 and 2019 that passed WGS QC criteria for analysis (Table 1). The median age of patients was 69 (range 30-87) and 24/25 (96%) were male. While this sex disparity is a limitation of our study in that potential sex differences may have been missed, it is in keeping with the disease burden seen in our practice in NSW, Australia, particularly for advanced and metastatic cSCC (Ashford et al., manuscript under review). This is in keeping with findings that age, male sex and immunosuppression are among the risk factors for metastasis [38]. Two patients were immunocompromised; one patient was on long-term azathioprine for rheumatoid arthritis and the other was on a combination of cyclophosphamide and tacrolimus following solid organ transplantation.

The location of the index primary lesion was known in 11 patients (Table 1). Nodal metastases were isolated from the neck in 13 patients and in the parotid in 12 patients. The majority of patients had either moderately differentiated (n = 8) or poorly differentiated (n = 12) cSCC, with evidence of extranodal extension found in 20/25 (80%) nodal samples.

### Tumor mutational burden (TMB)

Based on whole genome level calculations, the average TMB for SNVs and Indels across the 25 cases was 238.7 mutations per megabase (range 32.52 to 995.66 mutations/Mb) and 2.25 indel/megabase (range 0.63 to 5.9 mutations/Mb), respectively (Figure 1A, 1B; Supplementary Table 1) with the majority of somatic variants occurring in the non-coding regions as expected [12]. The only female tumor in this cohort had the second highest TMB at 499 mutations/Mb. There was no correlation between age, differentiation, nodal stage or extracapsular spread of the metastasis and TMB.

**Figure 1.**
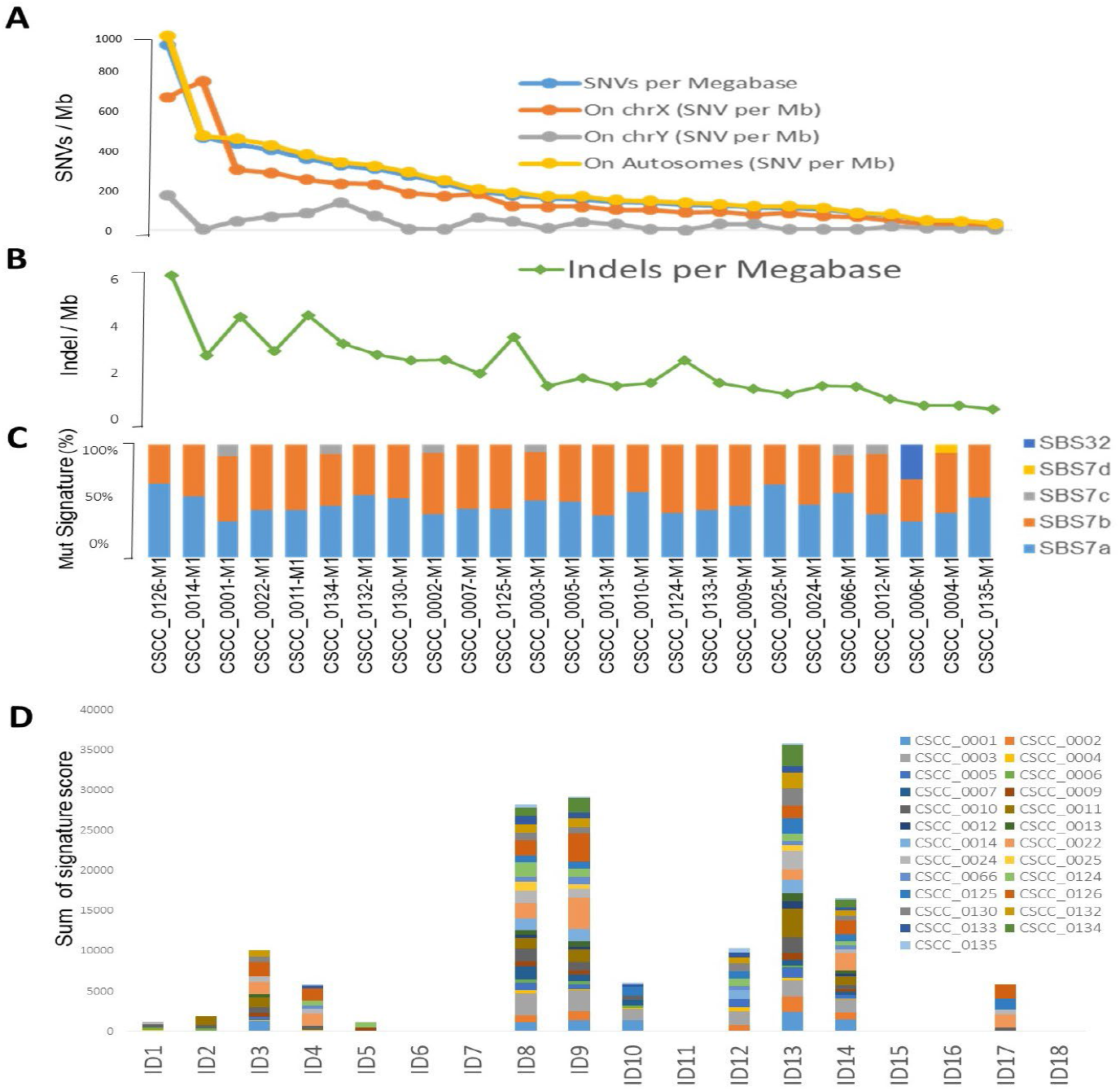
Overview of tumor mutational burden and signatures (whole genome-based). (A, B) illustrate the indel and SNV mutational burden in each sample, respectively. (C, D) show Indel (ID) and SNV mutational signatures for each sample, respectively, obtained using COSMIC V3.2 database. Full details are available in Supplementary Table 2 (Tabs 2-4).

### Mutational signatures

We performed mutational signature analyses of the 25 genomes based on COSMIC V.3.2 (https://cancer.sanger.ac.uk/signatures/). Signatures are designated as single base substitution (SBS), or small insertion and deletion (ID) signatures. SBS signatures 7a and 7b were the most prevalent (Figure 1C; Supplementary Table 2) in keeping with a UV association in metastatic cSCC as we previously reported in a smaller cohort using COSMIC V2 [12]. Substantial representation of SBS7c was also seen. SBS32 and SBS7d were observed in one sample. Indel signature analysis showed that ID8, 9 and 13 dominated over others (Figure 1D; Supplementary Table 2).

### Short variants

#### Coding Short Variants

The overwhelming majority of coding SNVs were missense mutations, followed by nonsense mutation, which represented less than 5% of variants (Figure 2A). Figure 2B shows various DNA sequence alterations, including single, double, and triple nucleotide variants as well as insertion and deletion (Supplementary Data 1). Over 80% of SNVs were C>T (Figure 2C, 2D). This is consistent with the dominant effect of UV radiation on pyrimidine bases and the UV signature referred to above and is independent of the degree of differentiation or any other clinicopathologic feature. Genes predicted to be driver genes via OncoDriveFML include *TP53, CDKN2A* and *ZNF730* having Q-values <0.1 (Figure 2E). MutSigCV and dNdScv analyses also found *TP53* and *CDKN2A* as the most significant mutated driver genes in our cohort (Supplementary Table 3). Genes that were predicted to be driver genes (P-value < 0.005) by at least two tools were considered for downstream analyses (Figure 2F). This resulted in 12 genes; *TP53, CDKN2A, C9, C9orf131, SLC22A6, KHDRBS2, COLEC12, LINGO2, CDHR5, ZNF442, PRLR*, and *DHRS4*. Of this list *TP53, CDKN2A* and *C9* were shared as significant by all 3 tools. Interrogation of the cBioPortal dataset for cSCC (metastatic = 92 and primary=88 cases) [17, 18, 20] with short variant analysis (Supplementary Figure 1) revealed recurrent mutations in *TP53, CDKN2A*, but also *C9, COLEC12* and *SLC22A6*. Not all genes identified as high impact and recurrent variants in our cohort were included in these targeted studies, which underscores the deficiencies of targeted analyses in discovery projects.

**Figure 2.**
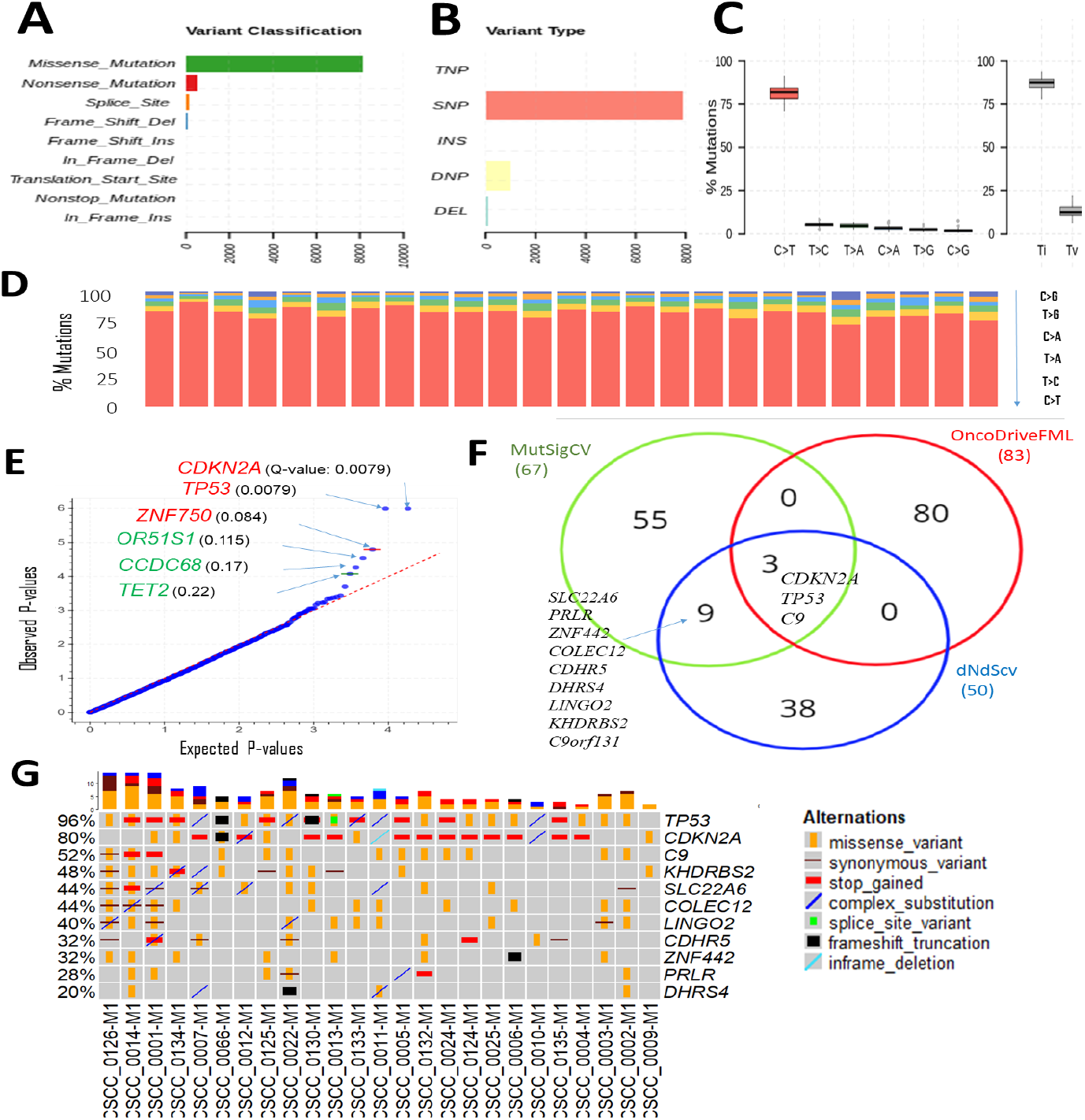
Overview of key coding mutations. (A) Variants classification, (B) variant types, (C) % transitions in all 25 samples and (D) % transitions for each sample. (E) Driver coding genes prediction results from OncodriveFML tool. The plot shows the most significantly altered genes (in the plots above the red line Q-values are below 0.1). Q-values are corrected P-values using the Benjamini/Hochberg correction (F) Venn diagram showing the overlap of genes predicted to be driver genes (P-value < 0.005) by 3 different driver detection tools, i.e. OncoDriveFML, MutSigCV and dNdScv. (For details refer to Supplementary Table 3). For further analysis, genes predicted to be driver genes by at least 2 tools were considered. (G) Detailed sample-level information of the SNVs along with types of variants in the top altered genes (from Figure 2F).

The only sample with no mutation in *TP53* was CSCC_0009 (Figure 2G). The TMB of this sample was 122/Mb, or 51% of the average across the cohort. Five samples without *CDKN2A* mutations averaged a TMB of 470/Mb, or 201% of the average for the cohort.

#### Variation in non-coding regulatory regions

The 3’UTRs that potentially play an important role in metastatic cSCC were discovered using OncodriveFML. SNVs within the 3’UTR region of *EVC, PPP1R1A, ABCA4*, and *LUM* showed significantly higher observed functional impact than the expected functional impact (Q-value

<0.03) (Figure 3A, Supplementary Table 3). We observed variation within the 3’UTR of both *EVC* and *PPP1R1A* in 48% of samples with a Q-value of 0.011 and 0.022, respectively (Figure 3B; Supplementary Table 4). The unique *PPP1R1A* variant with cDNA change of c.*491C>T [Chr12:54579896 (G to A)] was found in 5 samples (Supplementary Figure 2).

**Figure 3.**
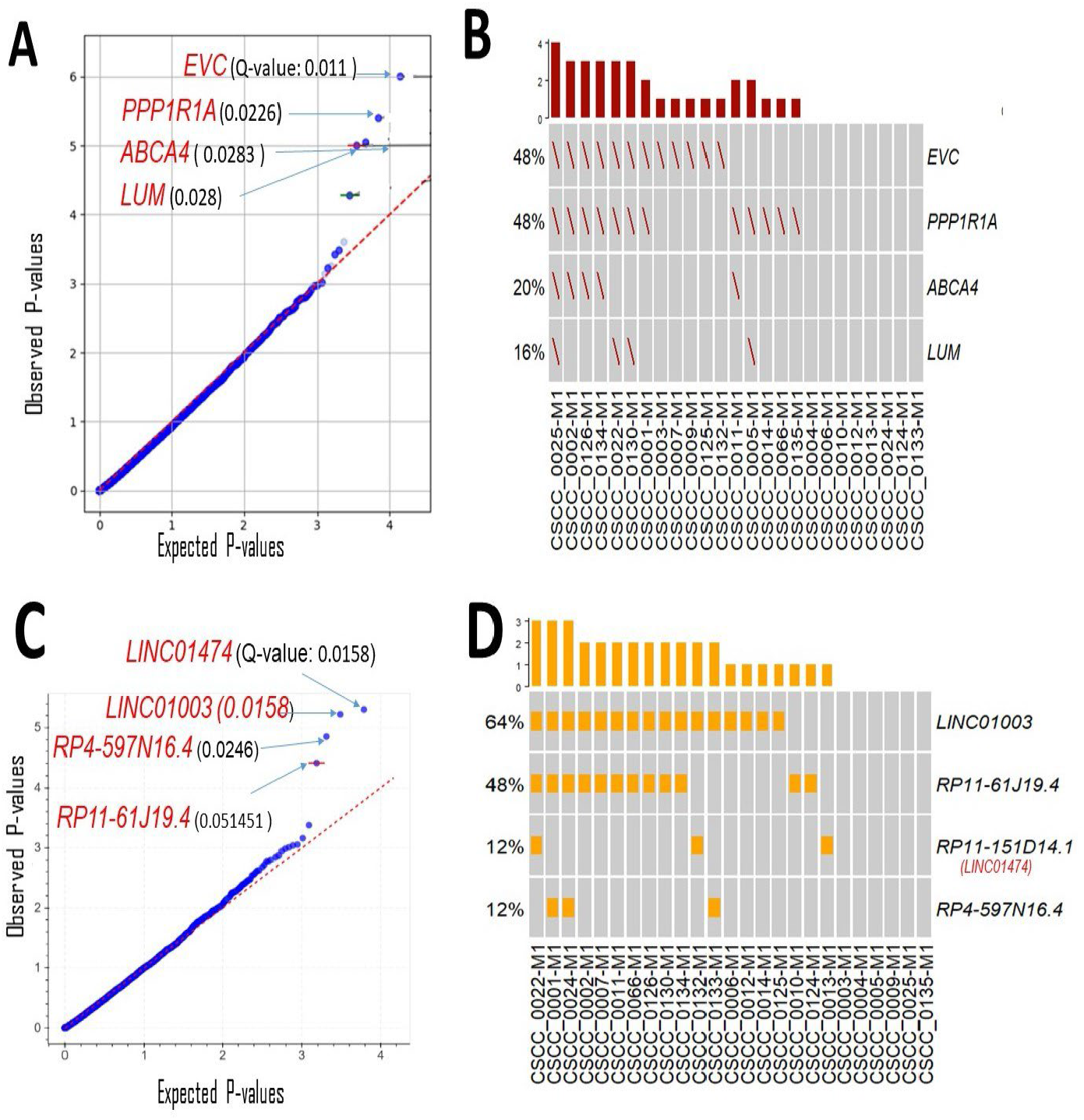
Driver genes prediction in non-coding genomic regions. Plots show the result of OncodrivFML (2.2.0) tool and mutations in the most significantly altered non-coding genes or regions in the cohort of 25 patient samples. (A) Potential 3’UTR regions associated driver candidates. (B) Variants with significantly altered 3’UTR regions. (C) Potential lncRNA driver’s candidates. (D) Variants with significantly altered lncRNAs. Plots in (A) and (C) show the frequency of observed mutations with respect to the expected frequency of the mutations in the corresponding regions. Q-values are corrected P-values using the Benjamini/Hochberg correction.

There are many reported limitations in the analysis and interpretation of 5’UTRs and promoters for high mutational burden tumors [39-41], a finding we also observed (Supplementary Figure 3).Currently no robust methodology exists to analyze these regions with confidence in cSCC thus analyses of 5’UTRs and promoter regions were not investigated further.

lncRNAs likely to have a potential impact on tumorigenesis were also predicted using OncodriveFML. Four lncRNAs were significantly (q < 0.05) biased towards high-impact mutations i.e *LINC01474* and *LINC01003, RP4-597N16*.*4*, and *RP11-61J19*.*4* (Figure 3C; Supplementary Table 3). Among these *LINC01474* and *LINC01003*, showed a high statistical significance Q-value of 0.0158. lncRNA *LINC01003* was altered in 64% of the cohort. Other recurrently mutated lncRNAs in our cohort was *RP11-61J19*.*4* (48% of samples) (Figure 3D; Supplementary Table 4).

### Structural and copy number variation

The extent of chromosomal copy number gain and loss was averaged across the genome for all 25 tumor samples (Figure 4A; Supplementary Table 5). Chr5p and 8q were the most frequently amplified regions, with 18q being the region with the most recurrent deletion. At sample level (Figure 4B), there were chromosome arm gains in Chromosome 7 and 5p in the majority of the samples and losses in 8p, 18q and 21q. Recurrent gain of 7, 8q, 5p and loss of 8p, 18, 21 was also previously reported by Pickering et al [20].

**Figure 4.**
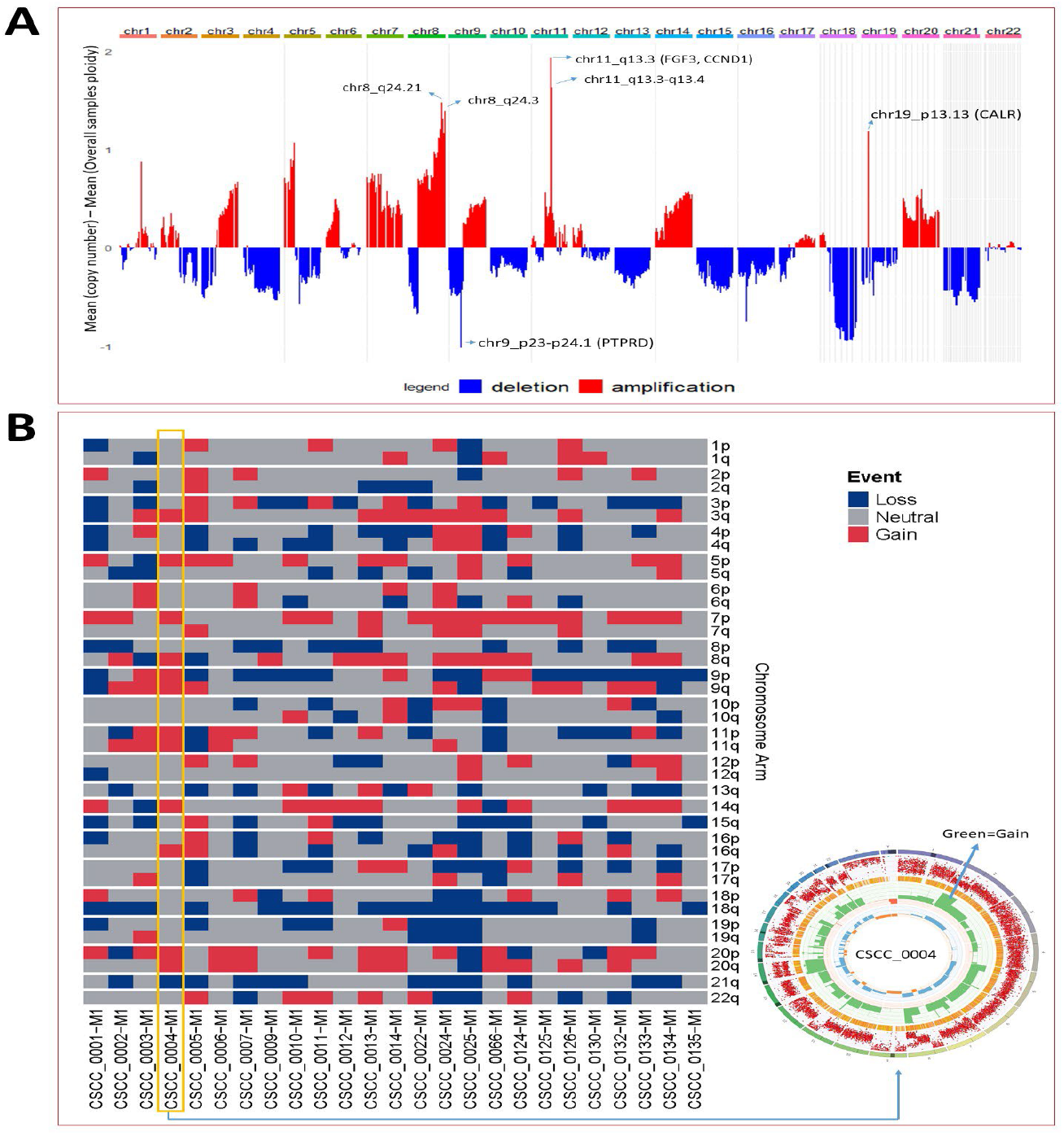
Chromosomal and recurrent genetic copy number variation. (A) Combined chromosomal CNV across 25 metastatic cSCC samples at the chromosomal level. The X-axis represents the differences of mean minimum copy number (bands) and means of overall samples ploidy (after adjustment for purity). Refer to Supplementary Table 5. (B) Chromosomes arm loss and gain at the sample level (Red denotes a gain, and blue denotes a loss). Both arms of chromosomes 7 and 5p show gains. 8p, 18q, and 21q show loss. (A chromosome arm is defined to be deleted if at least half of its bases are one or more copies less than the sample ploidy. A chromosome arm is defined to be amplified if at least half of its bases are one or more copies more than the sample ploidy.). Also shown is a Circos plot obtained from the PURPLE pipeline for CSCC_0004 as a representative example that summarizes various information at the sample level.

Structural variation analysis revealed that cSCC metastases are characterized by various complex, deleted, and unbalanced translocation events. Table 2 provides the summary of various structural events observed. Deletion and complex structural variants are common in cSCC; however, unbalanced translocation and other structural events were also observed (Table 2). The detailed effects of these structural events for putative oncogenes and tumor suppressor genes (TSG) are described in Table 3. Amplification events are linked to complex structural variants. Potential oncogene/TSG driver amplification and deletion were predicted by the PURPLE-GRIDSS-LINX pipeline, as reported in Table 3. Recurrent gene deletions were more common than gene amplifications. The most frequently deleted gene was *PTPRD (*Chr9p, 24% of samples). *PTPRD* deletion is already reported in primary and metastatic CSCC [42, 43]. Deletion of *PTPRD* (n=6) and *CDKN2A* (Chr9p) (n=1) did not co-occur in our cohort (Table 3), although *PTPRD* loss and significant mutation of *CDKN2A* co*-*occurred in 6 samples (CSCC_9, 11, 12, 133, 132 and 134) (Table 3 and Figure 2G). Deep deletion of *CDKN2A* was reportedin only 2/92 cases available on cBioPortal (Supplementary Figure 1).

**Table 2:** Summary of various event categories of structural variants. For more details, refer to Supplementary figures 4 and 5. Association can be noted between gain (Table 3) and complex SV events. The gene list was derived using LINX output. Only samples with events shown in Table.

| Sample | SGL | Del | DUP | Complex | UNBAL_Trans | Pair.Other | INF |
| --- | --- | --- | --- | --- | --- | --- | --- |
| CSCC_0001 | *SMAD4* | *SMAD4* |  |  |  |  |  |
| CSCC_0002 |  | *CDKN2A* |  |  |  |  |  |
| CSCC_0005 |  |  | *MYC* | *MYC* |  |  |  |
| CSCC_0007 |  |  |  | *CRLF2* |  |  |  |
| CSCC_0009 |  | *PTPRD* |  |  |  |  |  |
| CSCC_0011 |  | *PTPRD* |  | *CALR* | *HEBP2- NTRK2* |  |  |
| CSCC_0012 |  | *PTPRD* |  | *EGFR* |  | *PTPRD* |  |
| CSCC_0013 |  | *APC* |  |  |  |  |  |
| CSCC_0014 |  | *CREBBP* |  |  |  |  | *CREBBP* |
| CSCC_0025 |  | *CDKN2C* |  |  | *PARD6G* |  |  |
| CSCC_0066 |  | *PTPN13* |  |  |  |  |  |
| CSCC_0124 |  | *NEGR1* |  |  |  | *NEGR1* |  |
| CSCC_0132 |  | *PTPRD* |  | *RAF1-FGF3-CCND1* |  |  |  |
| CSCC_0133 | *PTPRD* | *PTPRD* |  | *CALR-chr1-chr3-chr6-chr8-chr22* |  |  |  |
| CSCC_0134 |  |  |  | *MCL1, CCND1-FGF3-Chr17* |  |  |  |
| CSCC_0135 |  | *PTPRD* |  |  |  |  |  |

**NBAL\_TRANS** -unbalanced translocation; **INF** = inferred breakend ; **DEL**=deletion; **DUP**=duplication; **SGL** =single breakend SV support

**Table 3:**
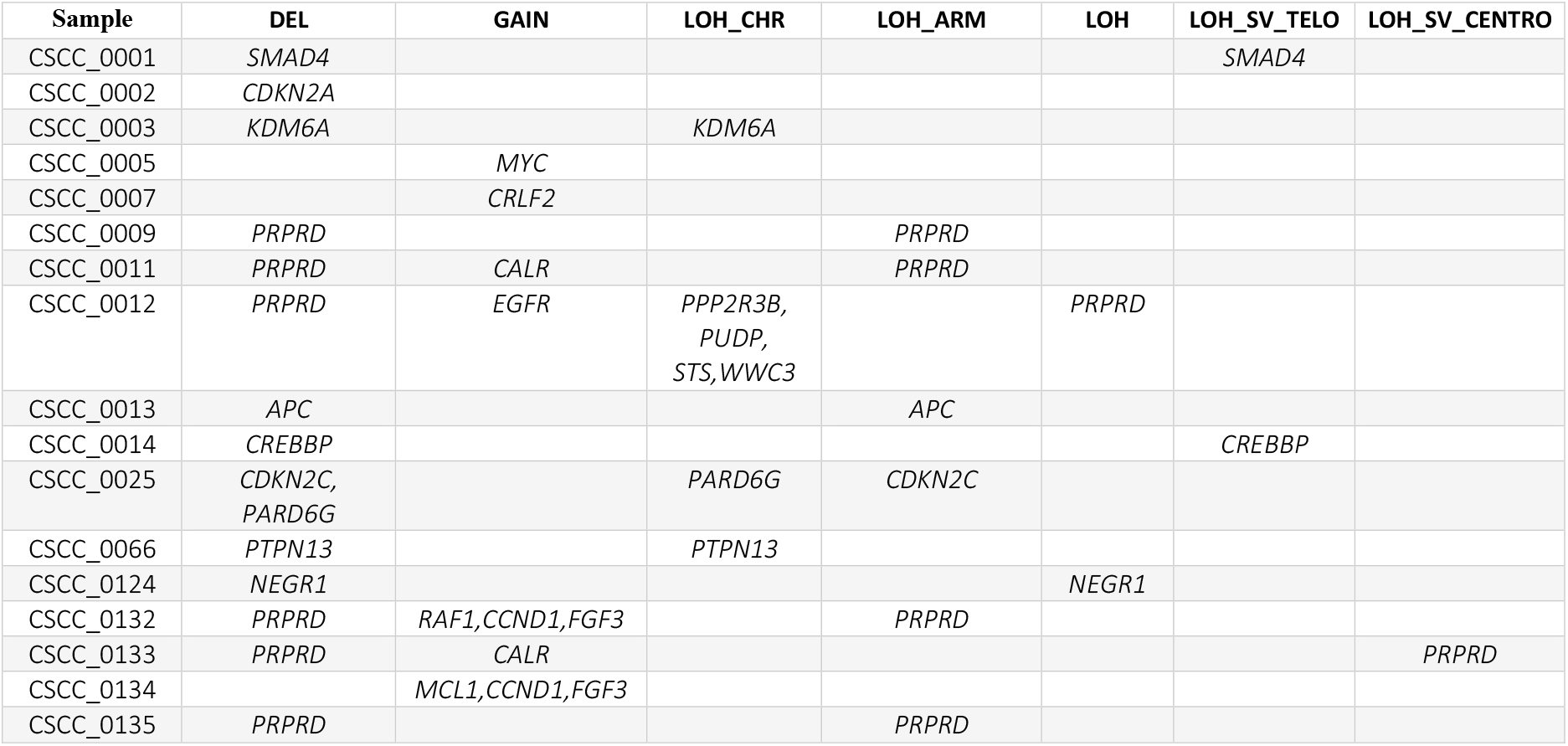
List of reportable drivers (Likelihood Type Onco/TSG) genes. Types of drivers are: GAIN = amplification by SV; DEL= homozygous deletion; LOH = focal LOH; LOH_ARM = chromosome arm level LOH; LOH_CHR = chromosome level LOH; LOH_SV_TELO = LOH from SV to telomere; LOH_SV_CENTRO = LOH from SV to centromere. Only samples with events shown in Table.

Loss of heterozygosity (LOH) was found at the focal, arm, chromosome, telomere, and centromere levels. The most common LOH events were that at the chromosome and arm level with these events concentrated to *PTPRD* locus (Table 3). No recurrent events for other genes among were observed (Table 3). Various examples of *PTPRD* structural events are reported in Supplementary Figure 4. A few other examples of the unbalanced translocation and complex structural variants are shown in Supplementary Figure 5.

The most frequently amplified genes (2/25, 8%) were *CALR, CCND1* and *FGF3* (Table 3). Interestingly *EGFR* was amplified in only one sample. Amplification of *CCDN1* and *FGF3* co-occurred in 2 samples (CSCC_0134 and CSCC_0132). *CCDN1* and *FGF3* are next to each other on the chromosome. These 2 cases had extensive nodal involvement (>50% of lymph nodes harboring tumor).

Despite this widespread genomic instability, only 2 coding-coding gene fusions were observed in our cohort. The first was between *STRN* and *DLG2* in sample CSCC_0009 (*STRN*: Exon 1 ENST00000263918 - *DLG2* Exon 7 ENST00000376104). *STRN* encodes a calcium-dependent calmodulin-binding protein [44]. *DLG2* plays a role in pain signalling and deletion is seen in both human and canine osteosarcoma [45]. We noted above that CSCC_0009 is the only sample without *TP53* mutations. CSCC_0009 came from a patient who had undergone liver transplantation and was on immunosuppressive therapy. The primary tumor that gave rise to this metastasis showed perineural involvement, which was also present in the metastatic deposit. The second gene fusion was between *NTRK2* and *HEBP2* in CSCC_0011-M1. This seems to be caused by unbalanced translocation event (**Supplementary Figure 5.B**).

### Enrichment analysis

Gene enrichment analysis was performed using the 21 genetically altered candidates identified above as significant/candidate driver genes, i.e., *TP53, CDKN2A, C9, KHDRBS2, SLC22A6, COLEC12, LINGO2, CDHR5, ZNF442, C9orf131, PRLR, DHRS4, PPP1R1A, EVC, LUM, ABCA4, LINC01003, LINC01474 (RP11-151D14*.*1), RP4-597N16*.*4, RP11-61J19*.*4*, and *PTPRD*.

The top significant pathway enrichment terms (Bio Planet 2019 [46]) are shown in Figure 5A. Most of the significant Bioplanet enriched terms come from *TP53* and *CDKN2A*, such as TP53 network, tumor suppressor ARF, CTCF pathway and cell cycle (G1/S checkpoint). However, *CDKN2A, LUM, CDHR5* and COLEC12 contribute to important cancer-related enrichment pathways, such as ‘TGF-beta regulation of extracellular matrix.’ Full details of these enrichment analyses are available in Supplementary Table 6.

**Figure 5.**
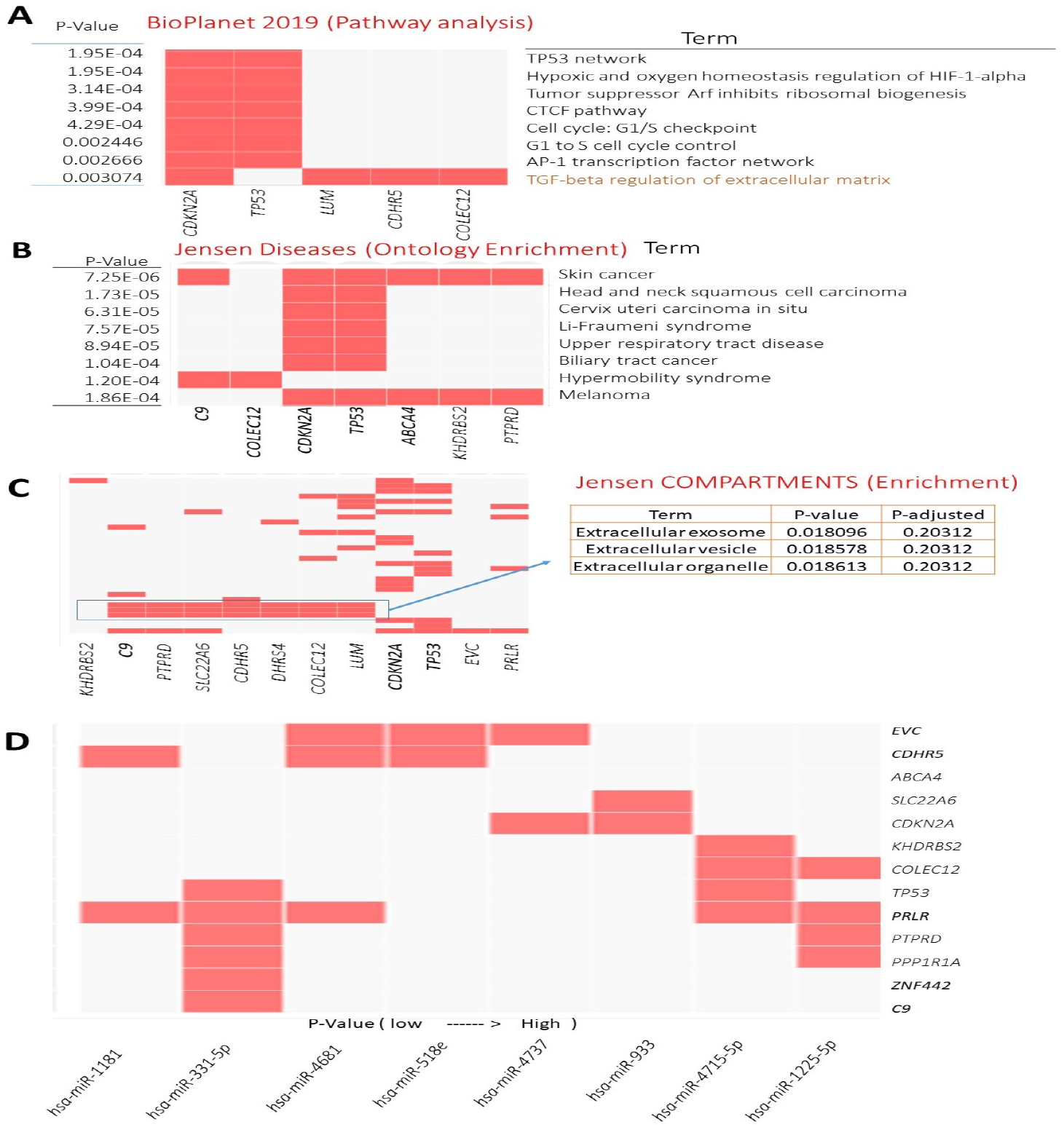
Enrichment analysis results of genetically mutated genes (21 candidates). (A) GO-Cellular Component terms showing 8 significantly enriched terms (obtained from BioPlanet 2019). (B) and (C) showing most significant Jensen diseases and Jensen compartments enriched terms, respectively. (D) Computationally predicted targets of miRNAs (TargetScan miRNA 2017). The x-axis represents the significance of the term. For details, refer to Supplementary Table 6.

The Jensen diseases enrichment tool identified skin cancer with highest significance (Figure 5B) with Jensen compartment-based enrichment analysis showing that most of these genes belongs to the extracellular compartment (Figure 5C). Other ontology enrichment analysis (MGI Mammalian Phenotype Level 4 2021; Supplementary Table 6) showed enrichment of increased fibroblast proliferation MP:0011703 where *CDKN2A, TP53 and LUM* alteration are the main contributors. We also predicted the miRNA targets for these driver candidates (Figure 5D). *hsa-miR-331-5p* was predicted to interact with 6 driver gene candidates, including *TP53* and *C9*. For this prediction, enricher platform use TargetScan miRNA database [47]. At the same time, hsa-miR-1181 was one of the most significantly enriched miRNAs for these driver candidates, however can target only 2 driver genes.

## DISCUSSION

This is the largest study to employ WGS to assess the mutational landscape of metastatic cSCC and demonstrates the breadth of somatic variation across non-coding and coding regions. Furthermore, we updated and expanded the understanding of UV-mutational signature patterns in metastatic cSCC [12], including the identification of novel Indel (ID) signature patterns. This highlights for the first time the nature and depth of variation within regulatory regions, with special attention devoted to UTR, and lncRNA. Additionally, we reported various structural events at whole genome scale for this diseases and also compared driver genes and SNVs to previous WES/targeted NGS studies on metastasis cSCC.

At 238 mutations/Mb (Median of 166.99 mutations/Mb) (at whole genome scale), the rate of TMB within metastatic cSCC is substantially higher than other cancers known to have high mutational burden, including melanoma (49 mutations/Mb) [48]. This finding is in keeping with Pickering et al [20] who found a median of 61.2 mutations/Mb from their WES of high risk primary (n= 32) and metastatic (n =7) cSCC being 4 times that of melanoma. The high TMB was associated with substantial structural variation, without recurrent gene fusions.

Alexandrov et al [49] detailed patterns of mutational signatures in 23829 tumor samples (1965 WGS) from the Pan Cancer Analysis of Whole Genomes (PCAWG) datasets including 17 small ID signatures, expanded to 18 in COSMIC version 3.2 (https://cancer.sanger.ac.uk) [50]. However, no cutaneous SCC (primary or metastatic) are included in this dataset. We identified the predominance of ID signatures 8, 9 and 13 (100% of samples effected) in our 25 metastatic cSCC samples. ID 8 is thought to be both related to double strand DNA break repair dysfunction and to age related changes. Melanoma is the only other cancer type reported to have a predominant ID 13 signature [49]. Our data also provides evidence of concomitance of ID 13 with SBS 7a and 7b (Figure 1C, 1D and Supplementary Table 2) in keeping with a UV-mediated mechanism for this signature. While we found ID 9 to be a dominant indel signature in cSCC it is rare in melanoma (2/104) but predominant in soft tissue sarcoma [49]. The mechanism of ID 9 is unclear but this departure from what is found in melanoma clearly shows some point of difference in these UV-induced skin cancers. When comparing the TMB associated with ID 9 signature among different cancers, the dominance in cSCC is clearly visible (Figure 6).

**Figure 6.**
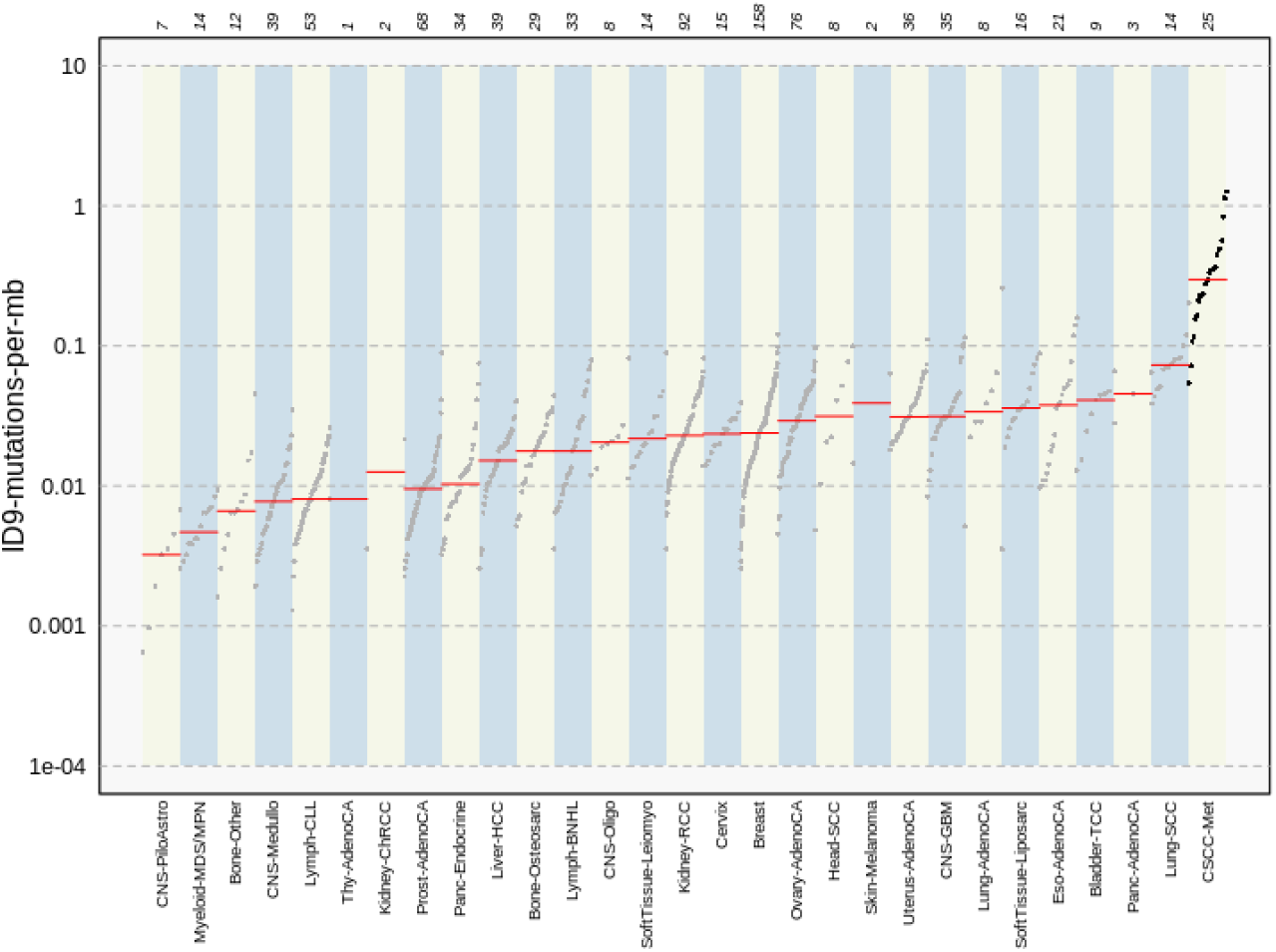
Comparison plot of ID9 mutations for various cancers. cSCC shows the highest ID9 mutations per Mb. The bottom x-axis represents the cancer types, and the upper x-axis shows the number of samples measured for specific cancer types. Y-axis indicates the number of mutations per Mb. Data for other cancers was obtained from ID9 signature details from COSMIC V3.2 and compared with cSCC data. cSCC data is calculated as ID9 signature score/3100 (Coverage for hg38 genome).

We identified substantial somatic variation within the 3’UTR region of *EVC, LUM* and *PPP1R1A. EVC* effects ciliary Hedgehog (Hh) regulation. Aberrant overexpression of *EVC* (and upregulation of Hh) has been reported in adult T-cell leukaemia as a result of epigenetic modulation [51]. The expression of *EVC* is reduced in nodal deposits of metastatic breast cancer compared with primary breast cancer suggesting a role in the metastatic process [52]. *PPP1R1A* is a protein phosphatase inhibitor which appears to have a variable but significant role in the metastatic process. For example, it is overexpressed in Ewing Sarcoma, and has been proposed as a driver of metastasis [53]. Conversely, levels of *PPP1R1A* were reduced in breast cancer when compared to adjacent non-diseased breast tissue [54]. Within our cohort, we observed a unique recurrent missense mutation in the 3’UTR of *PPP1R1A* in 5 samples.

*LINC01003* was the most mutated lncRNA in our cohort (64% of samples). In multiple myeloma, *LINC01003* behaves as a tumor suppressor genomic element. Up-regulation suppresses multiple myeloma by repressing cell viability and adhesion and promoting apoptosis. This effect is via its sponge effect on miR-33a-5p and its target *PIM1* [55].

As has been frequently reported for cSCC [5] (Supplementary Figure 1), *TP53* and *CDKN2A* were also the most recurrently altered genes in our cohort. Loss of function mutations within *TP53* and *CDKN2A* are well known to adversely impact cell cycle pathway control and DNA repair mechanisms. Kilnakis et al [56] describe a pattern of *TP53* mutation that differed between primary and metastatic disease in head and neck (mucosal) SCC. They found an overall lower rate of mutations in metastatic tumors, but a higher concentration of missense mutations in the DNA binding regions of the gene. However, Yilmaz et al [16] reported a significantly higher *TP53* mutation frequency in metastatic (85%) compared to primary tumors (corrected p-value <0.002). Additionally, they found a higher TMB with *TP53* mutation, but a worse response to immunotherapy. Burtness et al [57] reported that the extent of TMB in HPV-negative HNSCC is associated with loss of function mutations in both *TP53* and *CDKN2A*.

Of note in our study was the absence of significant or recurrent SNVs affecting *NOTCH1/2*. Inman et al [14] compared well differentiated to moderately and poorly differentiated primary cSCC and identified *NOTCH1, NOTCH2, TP53* and *CDKN2A* as the most commonly mutated genes, with *ATP1A1, HERC6, MAPK1P1L, GRHL2, TRAPPC9, FLNB* and *MAP3K9* identified as common early events in primary cSCC. Within this group, *GRHL2* was associated with less well differentiated tumors including those with a worse prognosis. In our cohort, only a single splice variant in *GRHL2* was identified, suggesting its role in metastatic disease is limited.

*C9*, (encodes complement component 9, *C9*) was also identified as a potential driver gene by 3 driver identification tools, with SNVs identified in 52% of the samples in our cohort. *C9* is a part of the membrane attack complex (MAC) and has been shown to modulate cellular behavior in the tumor microenvironment (TME) [58]. Since the TME plays a crucial role in tumorigenesis, progression, metastasis, and recurrence, *C9* might have significant potential in cSCC progression to metastasis. Various other components of the complement system have been linked to cSCC progression and immunosuppression and implicated as potential therapeutic targets [59-61]. With respect to *C9* specifically, it appears to be recurrently mutated in cSCC specimens (31% in primary and 10% in metastatic cSCC) as identified in the cBioPortal database (Supplementary Figure 1). and high expression levels have been proposed as a potential biomarker for the detection of gastric cancers [62] [63]. Further, the restrained expression of *C9* in tumor-associated macrophages promotes non-small cell lung cancer progression [64].

Apart from *TP53, CDKN2A* and *C9*, we identified 9 other potential driver genes with the most recurrently mutated gene being *KHDRBS2* (48% of cohort) with various impacts, including stop gained, complex and synonymous types apart from missense variant across the cohort. In the cBioPortal database, this gene is mutated in 20% of metastatic cSCC specimens (Supplementary Figure 1), suggesting it is a reasonably recurrently mutated gene in this disease.

Comparison of mutational frequency of primary and metastatic on the cBioPortal data suggests the potential of *COLEC12* (primary=25%; metastatic=60%) and *SLC22A6* (primary=16%; metastatic=30%) as a driver in metastatic cSCC (Supplementary Figure 1). Both *COLEC12* and *SLC33A6* are mutated in 44% of the samples in our cohort, and many of them are high impact SNVs. *COLEC12* is involved in leukocyte recruitment and cancer metastasis [65], and regulates the apoptosis of osteosarcoma [65]. Moreover, *COLEC12* is a potential biomarker of anaplastic thyroid cancer (ATC) [66]. In one cancerous study of gastric stromal cells (GSCs), the role of *COLEC12* is found in mediating the crosstalk between GSCs and dendritic cells (DCs) [67]. On the other hand, *SLC22A6* is known as an organic anion transporter 1 (*OAT1*). Expression and function alterations of *OAT1* play an essential role in therapeutic efficacy and the toxicity of many drugs. Such as for anti-cancer drugs methotrexate, Bleomycin, and Cisplatin-related toxicity [68-70]. *OAT1* variation associated with cardiotoxicity in pediatric acute lymphoblastic leukemia and osteosarcoma [71]. Furthermore, *OAT1* role in Breast cancer metastasis has been reported [72]. Important cancer-related roles of the other potential cSCC drivers are reported in Supplementary Table 7.

Loss of *PTPRD* was the most prominent copy number alteration in our 25 samples. *PTPRD* encodes protein tyrosine phosphatase receptor D, which belongs to a family of receptors whose action oppose that of the tyrosine kinases, which are central to cell growth and differentiation and oncogenic transformation. Large scale genomic events impacting *CDKN2A* can also affect *PTPRD* due to their proximity on chr9 [73]. In head and neck SCC, *PTPRD* inactivation significantly increases *STAT3* hyperactivation, which was associated with decreased survival and resistance to EGFR-targeted therapy [74]. *PTPRD* has been implicated as a tumor suppressor in several cancers with inactivating somatic variants found in >50% of GBM and between 10-20% of head and neck mucosal SCC (HNSCC) [75]. Lambert et al. [43] described deletions of *PTPRD* in 37% of metastatic primary cSCC and metastases. In addition, some of their cases also displayed a variant in the minor allele concordant with the deletion leading to a LOH event. It is thus possible that *PTPRD* plays a tumor suppressor role in preventing metastatic cSCC.

There were no recurrently amplified genes except for *CALR, CCND1* and *FGF3* which were each only amplified in 2/25 samples (Table 3). *CALR* encodes a ubiquitous endoplasmic reticulum bound calcium receptor [76]. Cellular stress can move *CALR* fragments to the plasma membrane from the ER and influence immune recognition of cancer cells. Recent analysis of *CALR* fragments in myeloproliferative disease suggest an immunosuppressive influence of extracellular *CALR* [77]. Cyclin D1 (*CCND1*) amplification is associated with nodal metastasis and worse survival in oral SCC [78]. In a review of *CCND1* copy number variation in metastatic non-cutaneous melanoma, amplification was prominent in those patients whose disease did not respond to immune checkpoint inhibition [79]. *FGF3* amplification is more common in metastatic breast cancer than primary tumors [80]. Targetable *FGF3* amplification was associated with a poorer prognosis and lung metastasis in hepatocellular carcinoma [81]. This amplification was seen in only 2% of total HCC but was most common in those cancers showing rapid response to sorafenib.

With respect to enrichment of driver gene alterations observed in our samples, dysregulation of the cell cycle pathway appears to be the central genomic theme of metastatic cSCC supported mainly by *TP53* and *CDKN2A. CDKN2A* encodes the CDK inhibitor p16^INK4a^. This inhibitor is an important controller of the activity of CDKs and progression from G1 to mitosis in the cell cycle. Inactivating mutations in *CDKN2A* with effects on p16^INK4a^ regulatory functions uncouple cell cycle control to promote tumorigenesis [82]. Interaction between *CDKN2A* and *TP53* through *MDM2* and its regulation by ARF (also encoded by *CDKN2A*) further disable cell cycle and apoptotic pathways (GO: Molecular function enrichment shows MDM2/MDM4 family protein binding).

The cellular process defined by the term “TGF beta regulation of extra cellular matrix” was also significantly enriched showing a role for *LUM, CDHR5, COLEC12* and *CDKN2A* in this process (Figure 5A). TGF-beta modulates the deposition of the extracellular matrix (ECM) and affects cell proliferation, differentiation, and migration.

Finally, miR-331–5p shows promise as a potentiator of cSCC drivers. miR-331-5p down-regulation contributes to chemotherapy resistance/relapse in leukemia [83] and it inhibits proliferation by targeting PI3K/Akt and ERK1/2 pathways in colorectal cancer [84].

## CONCLUSION

WGS provides insight into the unparalleled burden of mutation within metastatic cSCC, and our study has provided a deeper understanding of the genomic complexity of this disease. The functional impact of the varied and complex genetic alterations observed in metastatic cSCC should be validated in the future in confirmatory studies comparing whole genomes of non-metastatic primary tumours to metastatic tumours. This would significantly contribute to the identification of biomarkers in primary cSCC for predicting metastasis.

## Data Availability

All data produced in the present study are available upon reasonable request to the author

## ACKNOWLEDGMENTS

This work was funded by the Illawarra Cancer Carers, The Head and Neck Research Fund, Royal Prince Alfred Institute of Academic Surgery, The Cancer Institute NSW translational program grant, Chris O’Brien Lifehouse, National Health and Medical Research Council Project Grant APP1181179 and Tour de Cure. Authors would like to acknowledge National Computational Infrastructure (NCI-GADI) and Sydney Informatics Hub for computational services.

## AUTHOR CONTRIBUTIONS STATEMENT

AT performed the bioinformatics analyses and assisted in drafting the manuscript draft. BA conceived the idea, assisted in bioinformatics analyses and drafted the manuscript. DS performed bioinformatic analysis. MR assisted in drafting and editing the manuscript. BA and MR obtained funding for the project. JM and RG collated clinical data. JP, EM completed tissue processing. JP, EM, JC, RG, JL, SM reviewed and edited the manuscript.

## DATA AVAILABILITY STATEMENT

The data files used for genomic analysis have been deposited at the European Genome-Phenome Archive, which is hosted by the EMBL-European Bioinformatics Institute and the Center for Genomic Regulation, under accession number EGAS00001003370

