## Supplementary Figures for "Whole genome analysis reveals the genomic complexity in metastatic cutaneous squamous cell carcinoma"

**Supplementary Figure 1.** Oncoplot retrieved from cBioPortal comparing the variants of predicted driver genes determined by WGS of 25 metastatic cSCC genomes (refer to Figure 2). (A) Shows variants of metastatic cSCC studies submitted on cBioPortal (either whole exome or targeted panel based). *TP53* and/or *CDKN2A* alterations are present in all 92 metastatic samples. No alterations were reported for the other genes in 82 samples because of the use of targeted panel sequencing. (B) shows genetic alterations of 88 primary cSCC samples, where 56 cases are target sequencing based.

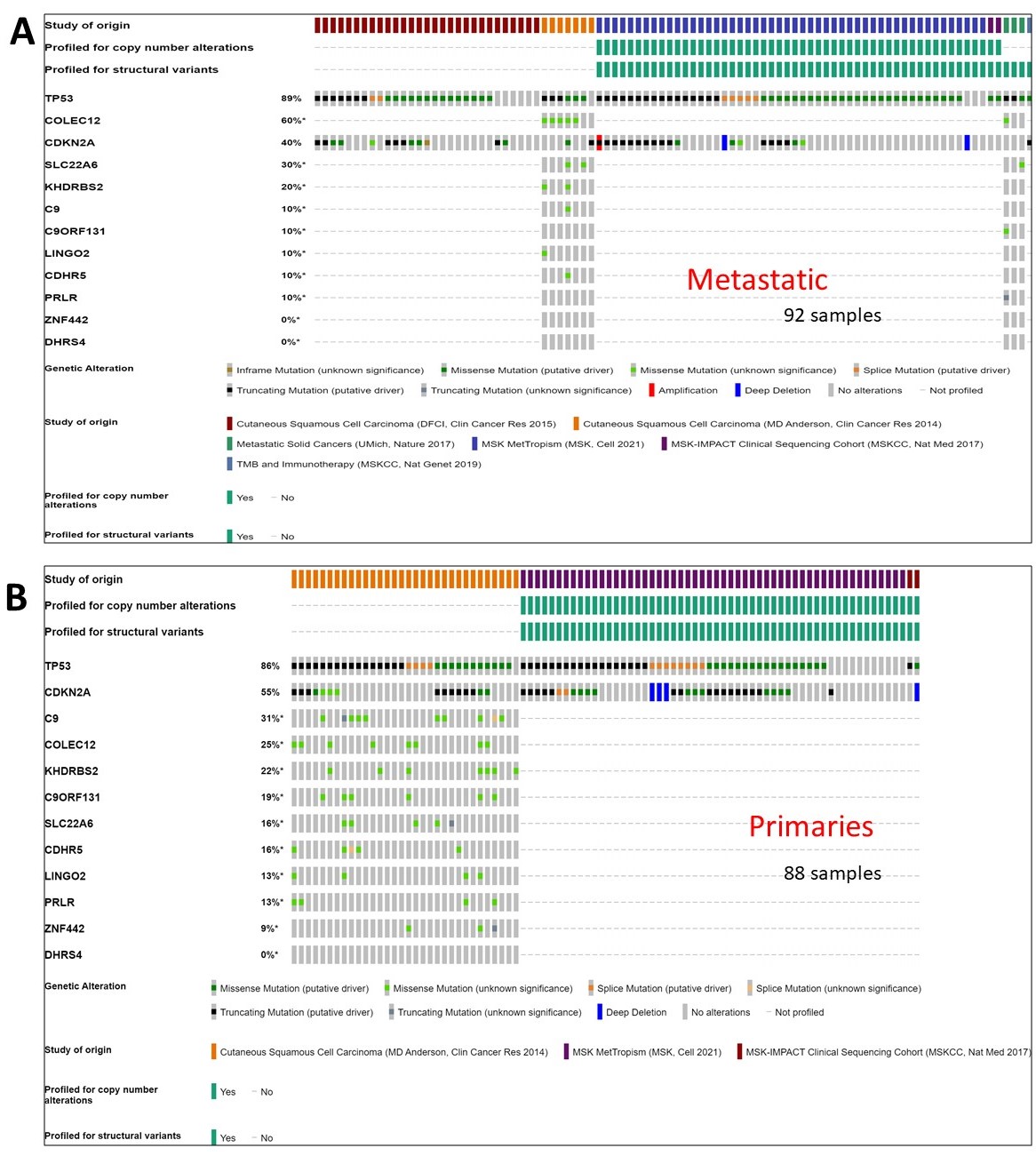

| Chrom | Position | Ref_Base | Alt_Base | Gene | Variant type | Samples |
| --- | --- | --- | --- | --- | --- | --- |
| Chr12 | 54579896 | G | A | PPP1R1A | 3’ UTR | CSCC_0001-M1;CSCC_0005-M1;CSCC_0011-M1;CSCC_0066-M1;CSCC_0126-M1 |

**Supplementary Figure 2.** IGV inspection. Examples of manual checks for recurrent variant in *PPP1R1A* (top) and *CDKN2A* (bottom) on IGV.

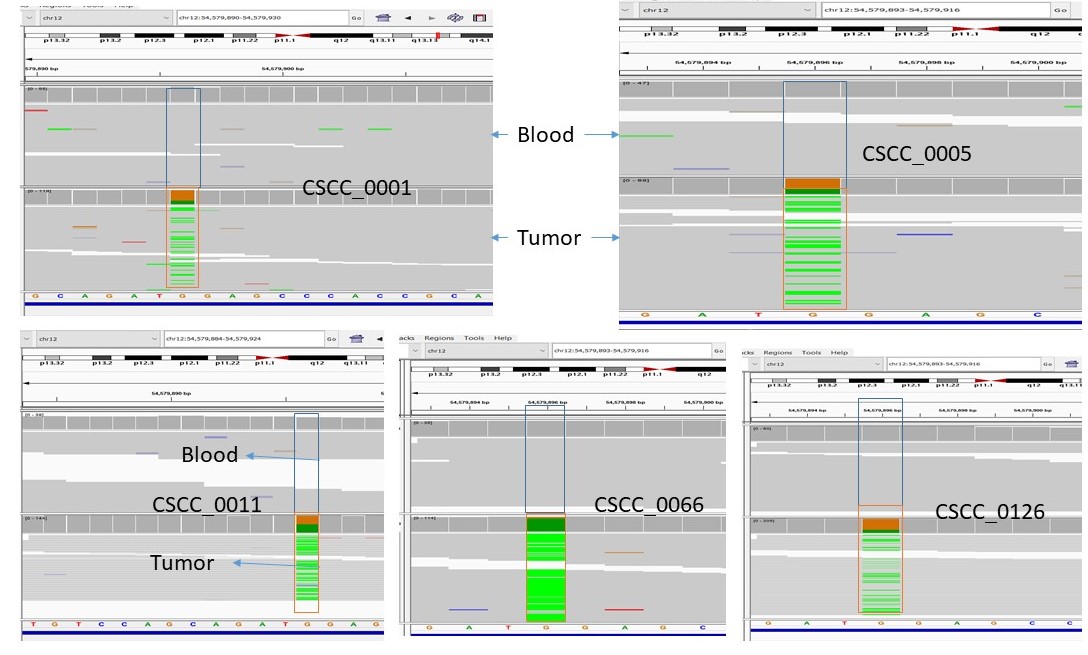

For *CDKN2A*

| Chrom | Position | Ref | Alt | Sequence_ontology |  | Protein_Change | Sample |
| --- | --- | --- | --- | --- | --- | --- | --- |
| chr9 | 21971134 | G | - | frameshift_truncation | c.225del | p.Ala76ProfsTer70 | CSCC_0066 |
| chr9 | 21971136 | G | A | missense_variant | c.223C>T | p.Pro75Ser | CSCC_0066 |

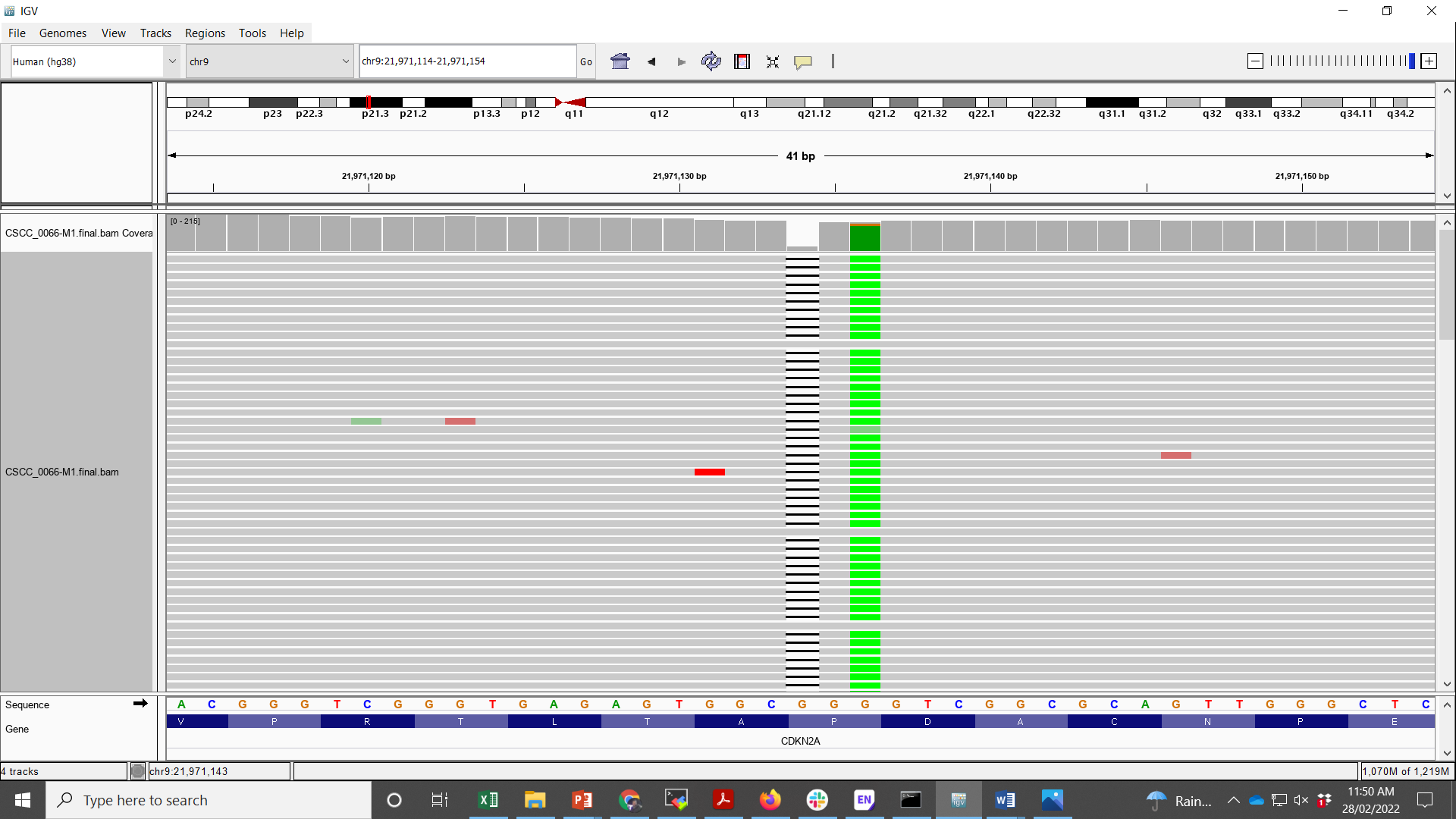

**Supplementary Figure 3**. Results obtained from OncodriveFML for 25 metastatic cSCC samples. The plots shown are similar to Q-Q plots with the Y axis showing the –log10 of the computed P-values (sorted) and the X axis showing the –log10 expected P-values (sorted). (A) 5’ UTR genomic element analysis, (B) Promoter region analysis. (A-B) Most genes are expected to follow the anticipated p-value trend in the figure, which is not the case for these regions (Due to abnormal accumulation of mutations). Since only a few % of genes follow the trends (which is just opposite to expectations), the genes listed are likely false-positive candidates.

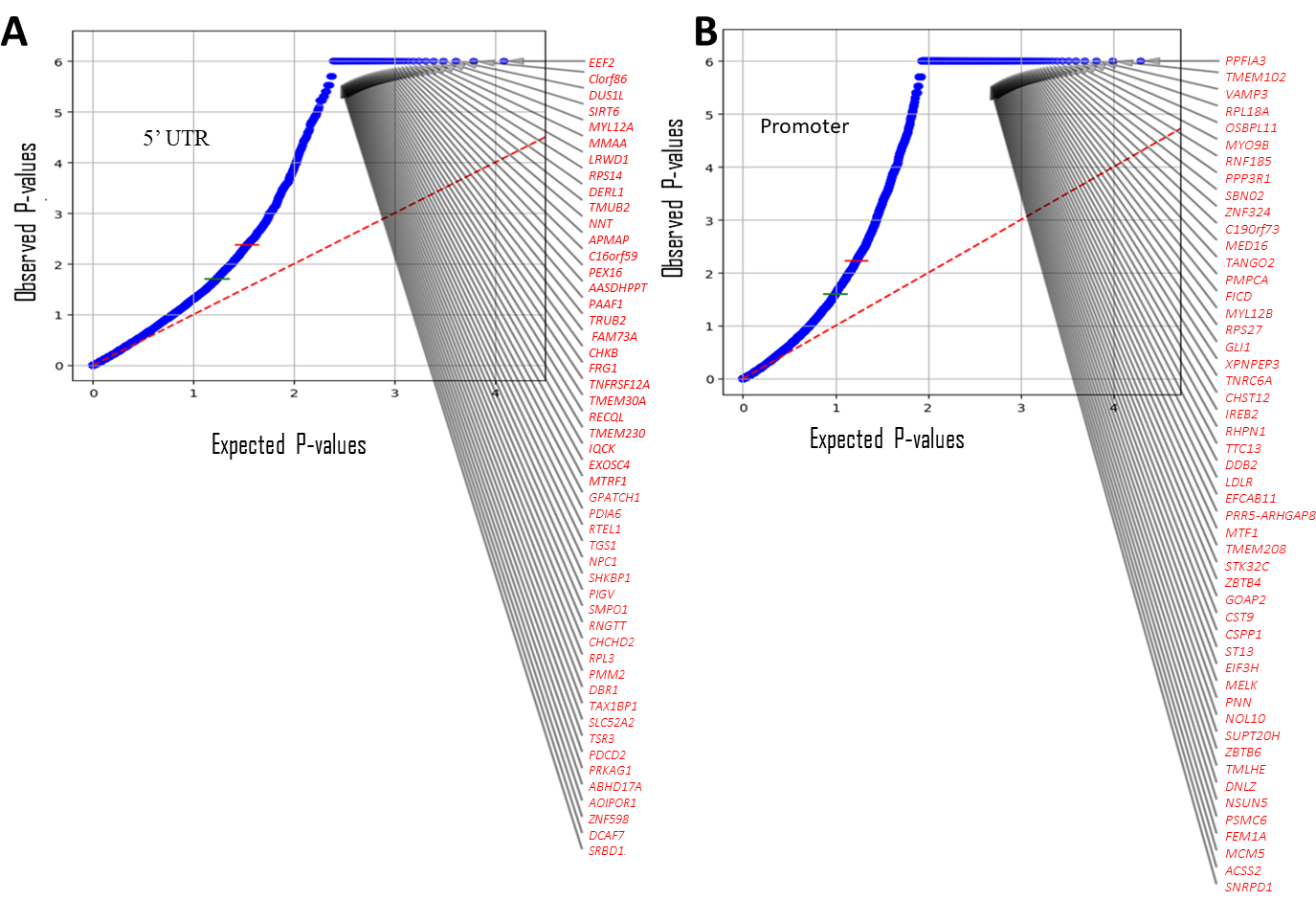

**Supplementary Figure 4.** Examples of structural variation in metastatic cSCC genomes for *PTPRD*. The top panel provides information regarding the interpretation of the Circos plots shown below (<https://github.com/hartwigmedical/hmftools/blob/master/linx/README_VIS.md>). Deletion structural Variants (Del SV) and single breakend SV support (SGL SV) are shown in the 4 Circos plots (CSCC_0009, CSCC_0011, CSCC_0132 and CSCC_0133) for *PTPRD*.

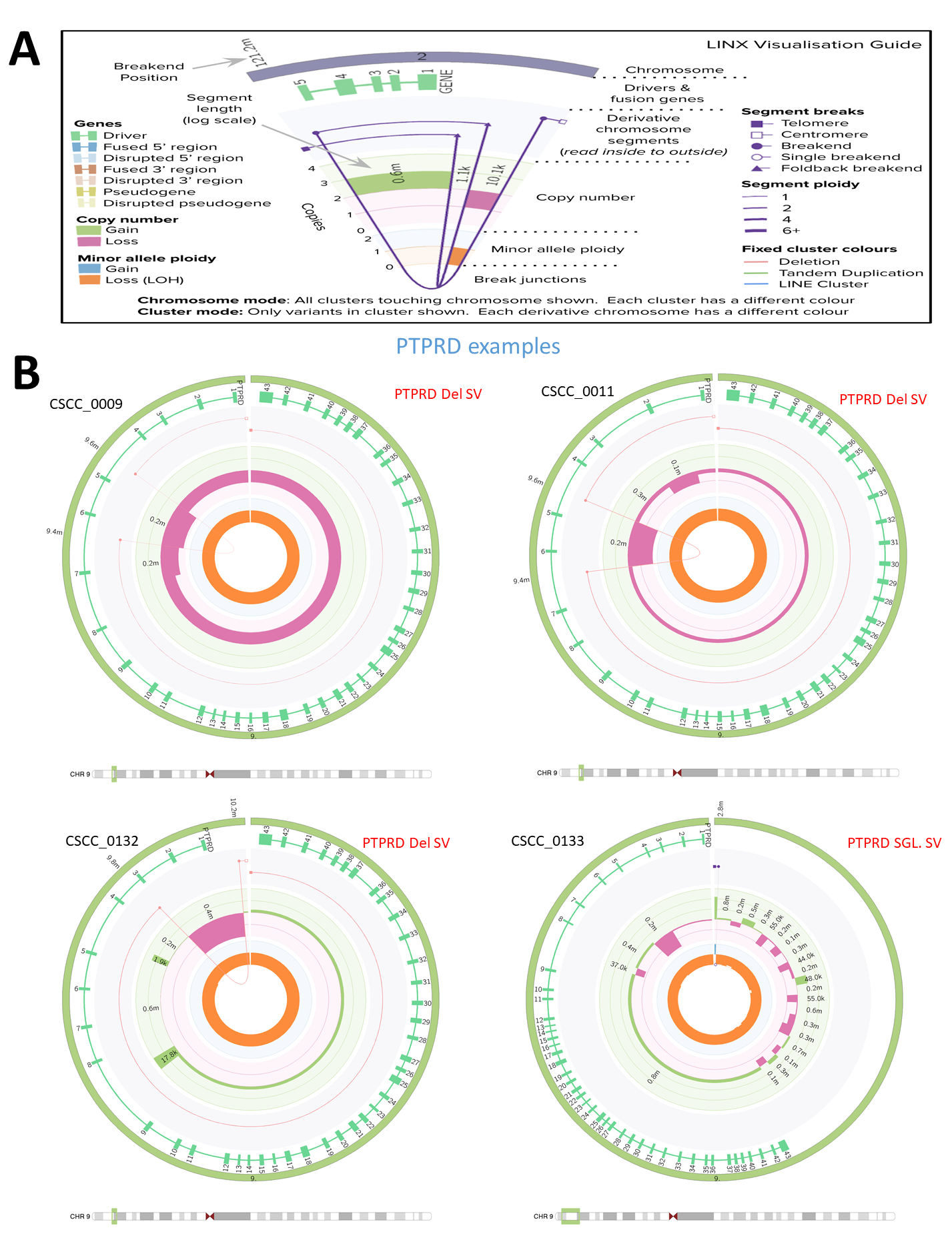

**Supplementary Figure 5**: Circos plot examples for other structural events (A) PARD6G unbalanced translocation (CSCC_0025). (B) In HEBP2- NTRK2 unbalanced translocation (CSCC_0011) event leads to gene fusion. (C) Complex Structural variant of CRLF2 (CSCC_007). (D) Complex Structural variant of RAF1-FGF3-CCND1 (CSCC_0132). For details, refer to <https://github.com/hartwigmedical/hmftools/blob/master/linx/README_VIS.md>

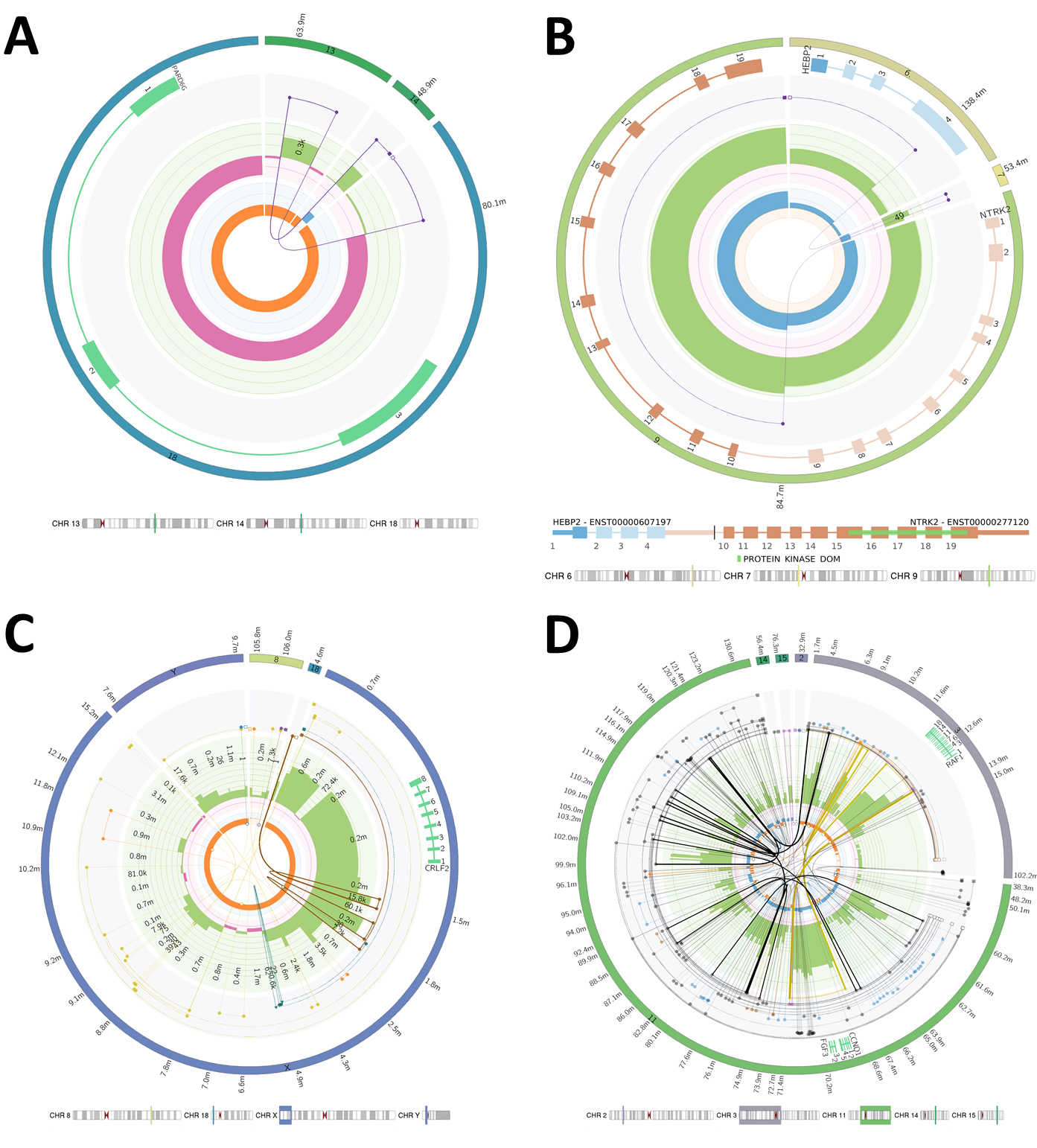
