## Supplementary Table 7 for "Whole genome analysis reveals the genomic complexity in metastatic cutaneous squamous cell carcinoma"

| **Gene** | **Role** | **Associated with** | **References** |
| --- | --- | --- | --- |
| KHDRBS2 | Carcinogenesis, alternating splicing in cancer | Aggressive renal cell carcinoma's, stage 4 neuroblastoma, survival in lung adenocarcinoma and Glioblastoma, papillary thyroid carcinomas | [1-5] |
| LINGO2 | Polymorphisms in the 9p21 region (*LINGO2*) are associated with the risk of multiple cancers, cellular proliferation and the development of features common to cancer cells. | Survival in advanced gastric cancers, interaction of LncRNA *LINC00518* and *LINGO2* acts as an Oncogene in Uveal Melanoma | [6-9] |
| CDHR5 | Encodes cadherin-related family member 5 | Prognostic marker of progression in clear cell renal cell carcinoma, inhibits proliferation of hepatocellular carcinoma, promotes malignant phenotype of pancreatic ductal adenocarcinoma | [10-12] |
| *ZNF422* | Specific to ethnicity or the tumor's heterogeneous populations | lung cancer (epigenetically silenced), homeostasis of MCF-7 breast cancer cells | [13-15] |
| PRLR | Carcinogenesis, GH signaling, EGFR/ERBB signaling pathways | metastatic risk in Breast cancer, carcinogenesis in cervical, ovarian, and endometrial cancers, a potential target for cancer treatment in prostate and human breast cancer | [16-20] |
| *DHRS2* | p53 regulation, cell cycle, apoptosis, and drug resistance in carcinoma cells | Motility in esophageal squamous cell carcinoma, poor prognosis in ovarian cancer, and a potential marker of breast cancer metastasis | [21-23] |

**Supplementary** **Table 7.** **Known** **role of other potential cSCC drivers in cancer.**

**References:-**
